# Differential Impact of Dyslipidemia on Chronic Kidney Disease between Men and Women

**DOI:** 10.1101/2022.06.22.22276767

**Authors:** Jay Dalal

## Abstract

Dyslipidemia (DLD), defined as an imbalance of lipids in the blood, is proven to have a strong association with chronic kidney disease (CKD). CKD is more frequent in women than men, although men are more likely to progress to severe stages. Whether sex differences drive the effect of DLD on CKD severity remains unclear. In this study, the relationship between biological sex and CKD severity was investigated by parsing the public electronic health records of 491 CKD patients in Abu Dhabi. Sex-specific relative risks (RRs) were calculated using a history of DLD as a risk factor for severe CKD (stages 3 - 5). The RR was 2.98 (95% CI: 1.78 - 4.18) in women and 2.86 (95 CI%: 2.02 - 3.70) in men. Smoking was identified as a confounding variable because it has a higher incidence in men and a known negative impact on CKD. Thus, we validated the findings in a subsetted group excluding any patient with a history of smoking: the RR was 2.96 (95% CI: 1.76 - 4.16) in women and 2.61 (95% CI: 1.57 - 3.65) in men. These results indicate that, among CKD patients, women with DLD are more likely to progress to severe stages of CKD than men with DLD. The excess risk may be explained by notably higher rates of obesity in women and the idea that women have poorer adherence to DLD medications, although this is newly discovered evidence. Future studies are needed to investigate mechanisms behind this observed disparity.

## Introduction

Chronic Kidney Disease (CKD) is defined by persistent renal structural abnormalities (e.g. cysts, tumors, atrophies, malformations, etc.) lasting longer than 3 months, leading to progressive loss of kidney function [1]. As a constantly increasing burden, due to considerable variety in risk factors, effect on 850 million people annually, and status as a strong risk factor for coronary artery disease [1], CKD presents a substantial clinical and public health issue. Therefore, developing a deeper knowledge about contributing factors to CKD pathogenesis and progression is instrumental to personalized care.

Dyslipidemia (DLD), defined as abnormally imbalance amounts of lipids, has a multifaceted but strong contribution to CKD development [2], mortality [1][2], progression to more severe stages and end-stage renal disease [3], and cardiovascular disease [3]. Lipids are absorbed from the intestines and used for a plethora of functions: energy, steroid production, bile formation, etc. DLD develops primarily due to increased triglyceride levels, decreased levels of high density lipoprotein (HDL-C), and varying levels of low density lipoprotein (LDL-C), all of which are factors critical to the pathways that absorb and utilize lipids [4]. The abnormal metabolism of these lipids and other CKD-related modifications of lipid particles promotes atherogenesis, inflammation, and endothelial cell dysfunction [1][6].

DLD plays an important role in understanding CKD physiology due to its high prevalence among DLD patients. In a 2002 study on more than a thousand patients on hemodialysis (a method that utilizes machinery to purify blood), only 20% had balanced and healthy lipid levels (LDL < 130 mg/dl, HDL > 40, triglycerides < 150). Additionally, of 317 patients on peritoneal dialysis (a method that utilizes the lining of the stomach as a natural filter to clean blood), 15% had normal lipid levels [12]. A larger study discovered that, out of more than 21,000 patients on dialysis, there was an 82% prevalence of dyslipidemia [12]. The National Kidney Foundation recommends that adolescents and adults with CKD be screened for DLD regularly using the standard fasting lipid profile, which measures total cholesterol, LDL-cholesterol, HDL-cholesterol, and triglycerides [6].

As CKD progresses, DLD often worsens: a National Health and Nutrition Examination Survey (NHANES) from 2001-2010 concluded that the prevalence of DLD increases from 45.5% in patients with CKD Stage 1 to 67.8% in patients with CKD Stage 4. Additionally, use of lipid-lowering medications, which treat DLD, increases from 18.1% in patients with CKD Stage 1 to 44.7% in patients with CKD Stage 4 [5]. Nevertheless, the true nature of DLD pathogenesis and its physiological effects on CKD remains unknown [6].

Numerous studies suggest that CKD is more prevalent in women than men, although men are more likely to progress to end-stage renal disease (ESRD) [7][8][9]. Nonetheless, clinically meaningful sex differences regarding the relationship between DLD and CKD, which could provide paradigm-changing insight on CKD-related DLD pathogenesis and management of DLD in patients with CKD, remain uncertain.

In this study, an analysis was conducted on an Abu Dhabian dataset to evaluate how links between a clinical history of DLD and severity of CKD differ between men and women [10].

## Methods

The electronic health records obtained for this study are from an open-access, public, and free dataset titled “Chronic Kidney Disease EHRs”. This dataset describes an observational study of 491 patients whose data was collected at Tawam Hospital in Al-Ain city (located in Abu Dhabi) between the 1st of January and 31st of December 2008. The cohort was made of 241 women and 250 men, with an average age of 53.2 years.

The data included 22 clinical variables, specifically demographics, physical conditions, a history of potentially relevant diseases, disease-specific medications, and results from clinical laboratory tests. According to the standards of Tawam Hospital, every patient included in the study was at risk for cardiovascular disease. Besides subsetting to make comparisons, no other manipulations were made to the data. Further information about the dataset can be found at the original article [10].

This study was conducted to assess how the impact of DLD in CKD progression differed between men and women. We used EventCKD35, a binary variable, to measure CKD severity. The estimated glomerular filtration rate (eGFR) baseline, which was collected by blood test and measures kidney function, was measured as a clinical variable and traditionally plays a key role in CKD staging definitions. An EventCKD35 value of 0 indicates that the CKD was at stage 1 (normal kidney function) or 2 (mild CKD), meaning that the true eGFR was ≥60. EventCKD35 of 1 indicated that CKD was at stage 3 (moderate CKD), 4 (severe CKD), or 5 (extreme CKD and kidney failure), meaning that the true eGFR was ≤59.

DLD was defined by a serum thyroglobulin (Tg) ≥ 2.26 mmol/L, a serum total cholesterol (TC) ≥ 6.21 mmol/L, or when the patient was taking lipid-lowering medications [10]. The sex variable was reported in binary format of male or female [10].

For the cohort presented, we calculated sex-specific Relative Risks (RRs) for patients with a history of DLD versus those with no history of DLD. Our defined risk factor was having a history of DLD, the positive outcome being an EventCKD35 of 0 and the negative outcome being an EventCKD35 of 1 (Table 1).

**Table 1.**
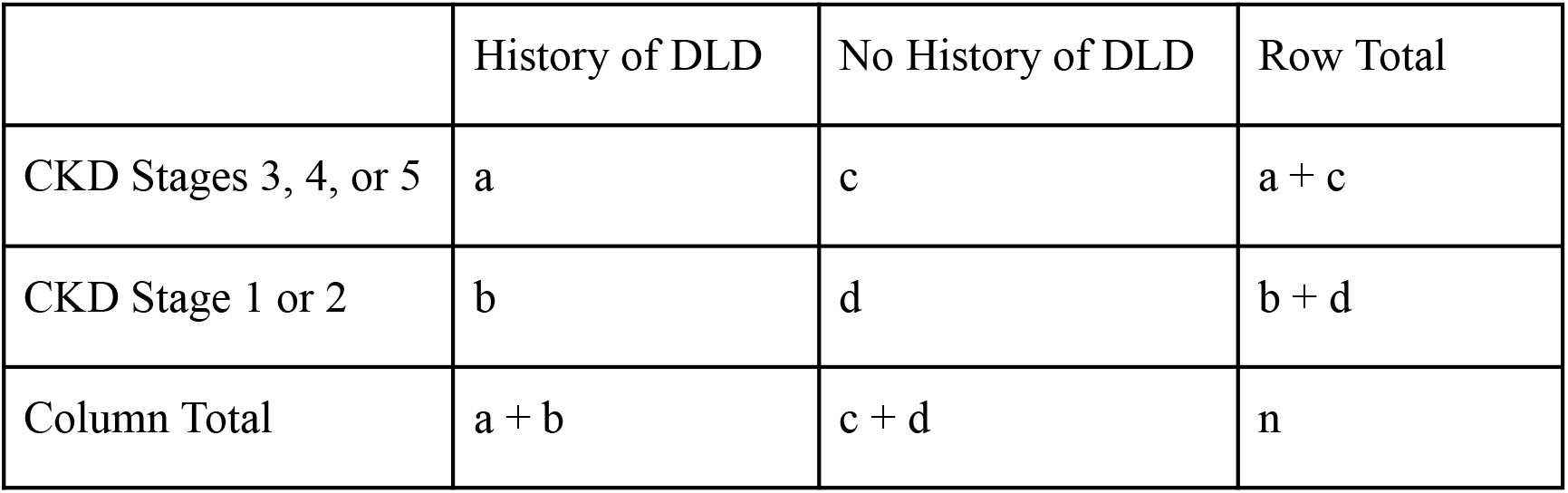
A 2×2 contingency table representing allocation of variables (a, b, c, d, and n) under which sex-specific calculations of Relative Risks (RRs) will be made.

Upon preliminary calculations on demographics and baseline characteristics of male and female patients, a history of smoking was identified as a confounding variable. In addition to worsening lipid imbalances and thus, increasing prevalence of dyslipidemia, including patients with a history of smoking, was proven to increase prevalence of other related conditions, such as coronary artery disease and hypertension [11]. Of the 75 patients with a history of smoking, there were 2 women (0.8% of all female patients) and 73 men (29.2% of male patients), presenting a wide sex-based disparity. For this reason, separate calculations for RRs for both a dataset with smokers and a subsetted dataset without smokers were performed to highlight the influence of a history of smoking on the results.

The Cochran-Mantel-Haenszel (CMH) test was performed to account for the binary nature of the predictor and outcome. Cases were stratified into two categories depending on sex. The strata-specific RR was calculated according to the equation (1), where a, b, c, d are the corresponding values in Table 1.

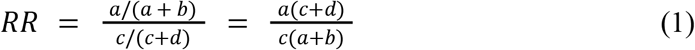

The weighted average across both the strata of male and female patients was calculated using equation (2) to provide a sex-independent relative risk for the entire patient population. This calculation was performed with the intention of providing a summarizing statistic of the dataset used in this study. This allows a comparison of results from this study to further studies examining the link between dyslipidemia and CKD.

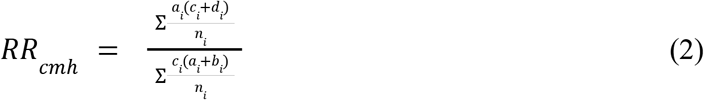

With *a*_*i*_, *b*_*i*_, *c*_*i*_, *d*_*i*_, and *n*_*i*_ representing the a, b, c, d, and n values, respectively, for the ith stratum (Table 1). The 95% Confidence Intervals (CI) were calculated according to equation (3)

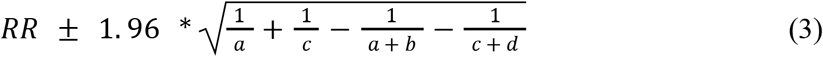

R version 4.2.0 was used to analyze the data.

## Results

Of the 491 cases in the original cohort, 416 were identified and had no reported history of smoking. Of the 75 smokers, many more were male (n = 73) than female (n = 2), presenting smoking as a confounding covariate. The median age for all patients was 53.0 years (SD: 13.82): 52.7 years (SD: 15.30) in males and 53.8 years (SD: 15.3) in females. Figure 2 includes the demographics, medical history, disease-specific medications, disease timings, and results of clinical/physical tests of all patients in the cohort, including smokers.

**Figure 2.**
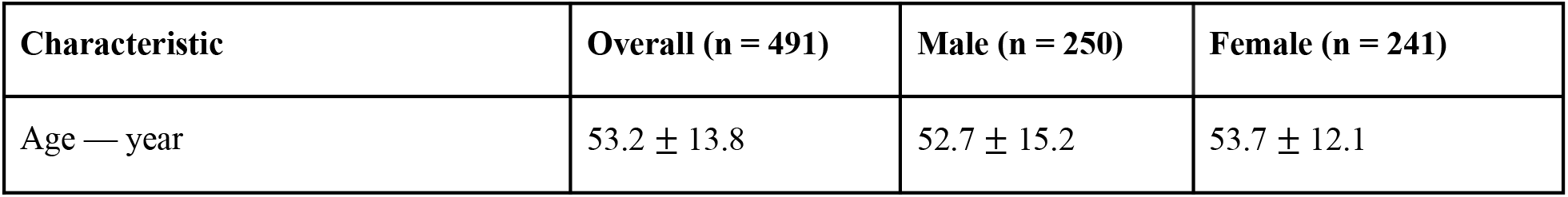
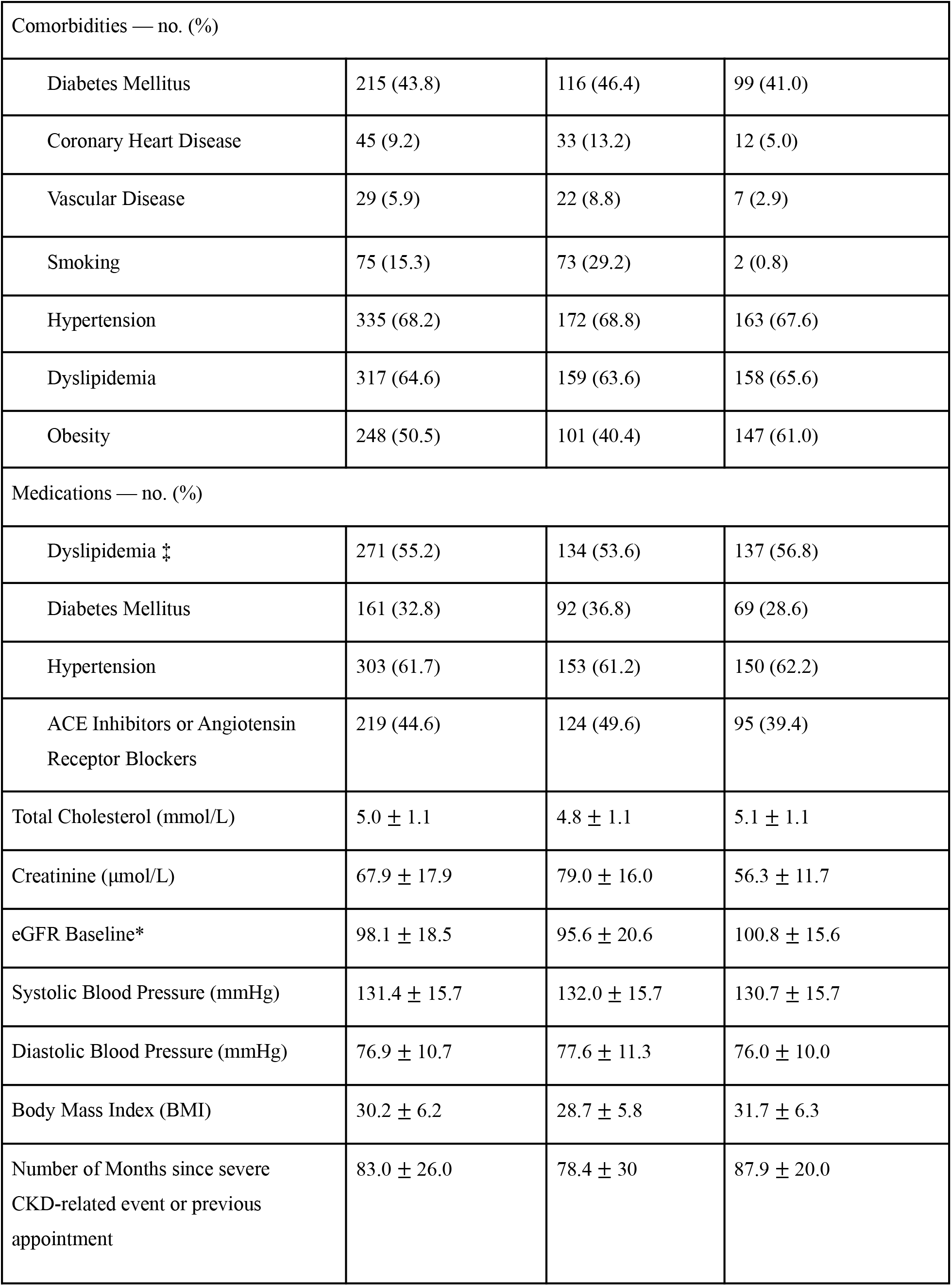
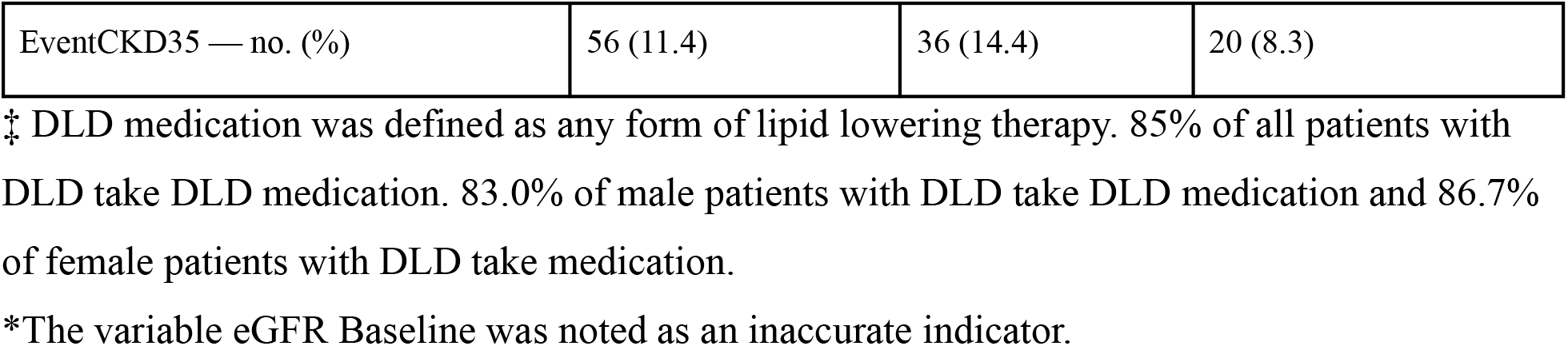
Baseline characteristics and results of clinical and physical tests for all patients in the cohort (n = 491). Numerical data is presented as mean ±standard deviation while binary data is presented as the number of positive cases with percent of positive cases in the total number of cases per category in parenthesis.

The RR of severe CKD in all women was 2.98 (n = 241, 95% CI: 1.78 - 4.18) while the RR of severe CKD in all men was 2.86 (n = 250, 95% CI: 2.02 - 3.70). The overall RR of all patients was 2.87 (n = 491, 95% CI: 2.18 - 3.56).

Subsetting the data to exclude patients with a history of smoking changed the results as follows: the RR of severe CKD in all non smoking women was 2.96 (n = 239, 95% CI: 1.76 - 4.16) while the RR of severe CKD in non smoking men was 2.61 (n = 177, 95% CI: 1.57 - 3.65). The overall RR of all non smoking patients was 2.73 (n = 416, 95% CI: 1.95 - 3.52) (Figure 3).

**Figure 3.**
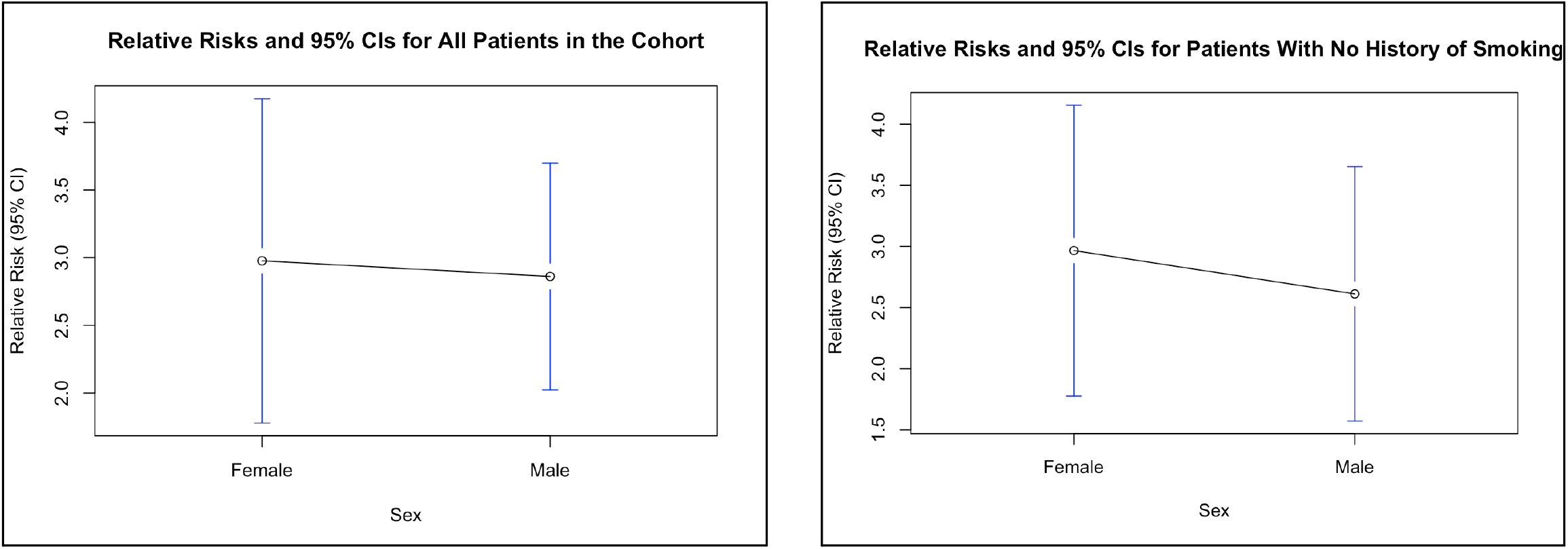
Summary of Calculated Relative Risks, represented by the dots, and 95% Confidence Intervals, represented by the dashes above and below the dots, by sex, showing both comparisons in the entire patient population and for the patient population without a history of smoking. Connecting lines are intended to visually demonstrate sex difference in relative risk.

## Discussion

In this retrospective cohort study, with data for 491 individuals with CKD in Abu Dhabi, DLD was stronger as a risk factor for CKD in women than men. For the entire population, women were found to only have 4.2% higher RR than men. Upon exclusion of any patient with a history of smoking, it was calculated that women had a 13.4% higher RR than men. This finding may have implications in developing a tailored strategy of care to prevent CKD progression in the patient population. For instance, personalizing treatment regimens for DLD in patients with CKD based on sex could potentially reduce this disparity.

Despite DLD’s importance in CKD progression, previous studies related to sex-related disparities address DLD as a risk factor for cardiovascular disease without accounting for it as a large risk factor for CKD due to cardiovascular disease’s higher prevalence and urgency. A recent study on 26,378 middle-aged rural Chinese residents concluded that, despite that fact that males were slightly older on average and more likely to have a non-physical job, smoke, consume more alcohol, have hypertension, have heartburn/regurgitation, and eat more unhealthy (spicy and fried) foods, women had a larger prevalence of dyslipidemia and more unbalanced lipids. Women however, did have higher rates of obesity and a history of diabetes [14].

Consistent with this finding, our study found that men followed unhealthier habits than women in every category except for a few: women have a slightly higher BMI and cholesterol, and were 1.5 times or 19.7% more likely to have obesity (Fig. 2). Obesity has been continuously proven to be a large promoter of CKD progression. A high BMI causes increase in intraglomerular pressure, leading to nephron damage that is often permanent. Obesity-related glomerulopathy, a condition marked by damage of the renal glomerulus, has been an exponentially more common condition in recent years [15]. Regardless, this obesity would usually be borderline as the definition of obesity in the United Arab Emirates is a BMI > 30 [16] and women had an average BMI of 31.7 while men had an average BMI of 28.7. Thus, this may not fully explain the disparity of CKD progression.

Another possibility is that women tend to have a higher difficulty relating to DLD medication adherence. The study for the dataset defines DLD medications as any lipid lowering therapies, of which statins are the most commonly used. A review on sex differences of statin adherence observed that in both intentional and unintentional statin adherence, women were less likely to adhere due to decreased healthcare provider awareness surrounding women with DLD, physiology differences, and traditional family responsibilities [17]. It is important to note that this review addresses statin usage for reducing risk of cardiovascular disease and that evidence concerning statin usage to treat DLD and prevent cardiovascular disease in CKD patients is conflicting and inconclusive [6][18]. Interestingly, in our dataset, there is a slightly higher percentage of women taking DLD medication (out of women with DLD) than men taking DLD medication (out of men with DLD).

This study came with numerous strengths. The fact that we analyzed a specific dataset allowed us to have access to the electronic health records of specific patients. Additionally, the identification of smoking as a confounding variable due to the fact that smoking was much higher in prevalence in men than women allowed us to both further prove smoking as a factor that promotes CKD progression while adjusting our study so that our findings can be applicable to regions where culture around smoking is different.

Although this analysis provided valuable insight into personalized care for DLD pathogenesis in CKD, it had some limitations. Due to the minimal amount of sex-specific and DLD-specific CKD data that is online and accessible, the dataset that we used was small in size, with only 491 patients, and involved data collection in a single hospital over a single year. This may increase the likelihood that our findings don’t include CKD patients from countries and cities in different regions with different cultural and racial characteristics. The dataset also didn’t include properly collected data on eGFR baseline and, as a result, we had to utilize the Event CKD35 variable to make comparisons between mild CKD and moderate to severe CKD. Using accurately measured eGFR baseline, we could have provided a more valuable and sophisticated insight into differences of DLD related pathogenesis between the sexes. Another limitation in the dataset was that it involved data collected at a single point in time. Studying DLD pathogenesis over multiple points in time may provide further insight on progression, rather than just severity.

## Conclusion

This study demonstrated an unequal impact of DLD between women and men on CKD severity, with women having the disadvantage when it comes to severity. This disparity is unlikely to be solely explained by the higher rate of obesity among women and future studies should explore sex differences with statin adherence in CKD. Future studies could explore sex differences of DLD in CKD using meta-analyses and other statistical tools, analyze the data from the most refined measurement of eGFR, and study this phenomenon in other regions, especially more ethnically and racially heterogeneous ones.

## Data Availability

All data produces are available online at: https://www.kaggle.com/datasets/davidechicco/chronic-kidney-disease-ehrs-abu-dhabi.

## Acknowledgements

This paper would not have been possible without the mentorship of Laken Rivet through Polygence. I also thank Courtney Smith for providing her comments on this manuscript before submission.

